# Stillbirth rates and their determinants in a national maternity hospital in Phnom Penh, Cambodia in 2017-2020: a cross-sectional assessment with a nested case-control study

**DOI:** 10.1101/2023.08.29.23294731

**Authors:** Aliki Christou, Jackline Mbishi, Mitsuaki Matsui, Lenka Beňová, Kim Rattana, Ayako Numazawa, Azusa Iwamoto, Sophearith Sokhan, Nary Ieng, Thérèse Delvaux

**Affiliations:** Department of Public Health, Institute of Tropical Medicine, Antwerp, Belgium; Hasselt University, Hasselt, Belgium; Department of Public Health, Kobe University Graduate School of Health Sciences, Japan; Nagasaki University School of Tropical Medicine and Global Health, Nagasaki, Japan; National Maternal and Child Health Center, Phnom Penh, Cambodia; Bureau of International Health Cooperation, National Center for Global Health and Medicine, Japan

**Keywords:** stillbirth, fetal death, risk factors, Cambodia, case-control study, maternal health, perinatal health, routine data, obstetric emergency, hospital, referral

## Abstract

**Background:** In Cambodia, stillbirths and their underlying factors have not been systematically studied. This study aimed to assess the proportion and trends in stillbirths between 2017-2020 in a large maternity referral hospital in the country and identify their key determinants to inform future prevention efforts.

**Methods:** This was a retrospective cross-sectional analysis with a nested case-control study of women giving birth at the National Maternal and Child Health Centre (NMCHC) in Phnom Penh, 2017-2020. We calculated percentages of singleton births at ≥22 weeks’ gestation resulting in stillbirth and annual stillbirth rates by timing: intrapartum (fresh) or antepartum (macerated). Multivariable logistic regression was used to explore factors associated with stillbirth, where cases were all women who gave birth to a singleton stillborn baby in the four-year period. One singleton live birth immediately following each case served as an unmatched control. Multiple imputation was used to handle missing data for gestational age.

**Results:** Between 2017 and 2020, 3.2% of singleton births ended in stillbirth (938/29,742). The stillbirth rate increased from 24.8 per 1000 births in 2017 to 38.1 per 1000 births in 2020, largely due to an increase in intrapartum stillbirth rates which rose from 18.8 to 27.4 per 1000 births in the same period. The case-control study included 938 cases (stillbirth) and 938 controls (livebirths). Factors independently associated with stillbirth were maternal age ≥35 years compared to <20 years (aOR:1.82, 95%CI:1.39,2.38), extreme (aOR:3.29, 95%CI:2.37,4.55) or moderate (aOR:2.45, 95%CI:1.74,3.46) prematurity compared with full term, and small-for-gestational age (SGA)(aOR:2.32, 1.71,3.14) compared to average size-for-age. Breech/transverse births had nearly four times greater odds of stillbirth (aOR:3.84, 95%CI:2.78,5.29), while caesarean section reduced the odds by half compared with vaginal birth (aOR:0.50, 95%CI:0.39,0.64). A history of abnormal vaginal discharge increased odds of stillbirth (aOR:1.42, 95%CI:1.11,1.81) as did a history of stillbirth (aOR:3.08, 95%CI:1.5,6.5).

**Conclusions:** Stillbirth prevention in this maternity referral hospital in Cambodia requires strengthening preterm birth detection and management of SGA, intrapartum care, monitoring women with stillbirth history, management of breech births, and further investigation of high-risk referral cases.

**Plain English summary:** In Cambodia, there is very little information published on stillbirths to know precisely how many there are and to understand the underlying reasons they occur so they can be prevented in the future. Our study aimed to quantify the number of stillborn babies and identify some underlying risk factors from one of the largest maternity referral hospitals in the Phnom Penh, Cambodia. We examined data from almost 30,0000 health facility medical files of women who gave birth between 2017 and 2020 which included 938 stillbirths. We found that about 3.2% of births ended in a stillbirth and that this percentage increased between 2017 and 2020. Women who had preterm babies, or whose babies were small in weight for their gestational age, and babies that were born breech had a higher chance of being stillborn. Women who had abnormal vaginal discharge which can indicate a possible infection also had a higher odds of having a stillbirth. We also found that women who had a stillbirth previously had almost three times higher chance of having another stillborn baby. Having a caesarean section reduced the likelihood of having a stillborn baby by about half. These findings suggest that efforts are needed to better identify and manage women with preterm births and monitor fetal growth as well as ensure breech births are managed adequately.

## Background

Stillbirths comprised 36% of deaths of children under-five years globally in 2021 and their contribution to these deaths has increased from 23% in 2000.[1] Of the estimated 1.9 million stillbirths worldwide every year, almost 90% occurring in low or lower middle-income countries.[2] The 2014 global commitment to The Every Newborn Action Plan set a stillbirth rate target of 12 per 1000 births for all countries to reach by 2030.[3] Yet declines in stillbirths globally have been slow and are not on track to reach this target. The highest stillbirth rates are in Sub-Saharan Africa at 21 stillbirths per 1000 births followed by South Asia at 17 per 1000 births.[2] The absence of quality data in routine health information systems has limited our understanding of the true burden and precise causes and risk factors for stillbirths at country and sub-national levels, and contributes to the lack of prioritisation of policy and health interventions to reduce stillbirths.[4]

The stillbirth rate is defined as the number of babies born without signs of life from either 22 weeks or 28 weeks’ gestation per 1000 total births with the latter recommended for international comparisons[4, 5]. The stillbirth rate is an important indicator of the quality of care received during pregnancy and childbirth.[6] Most stillbirths can be prevented if access to high quality of care can be ensured throughout the maternal continuum of care.[7] Factors known to be associated with stillbirth include complications in childbirth, maternal conditions such as hypertension and diabetes, infections including syphilis, HIV and group B streptococcus, haemorrhage, and genetic conditions.[7] The contribution of these causes and risk factors can vary across different contexts and data is needed for each context to prioritise and target preventive strategies.

In Cambodia, national stillbirth rates have been estimated from modelling and suggest an impressive decline by nearly half from 24.7 per 1000 births in 2000 to 11.4 per 1000 births in 2021[2], indicating that the country already reached the 2030 target. However, these national rates derived from mathematical estimations can have wide uncertainty intervals and mask within-country variability. Cambodia has almost universal coverage of facility births with 98% of women giving birth in a health facility in 2020[8], thus, examining routine facility data is an important data source which can provide insights into key contributing factors without a high risk of selection bias. Despite this high coverage, there are few published studies investigating risk factors for stillbirth in Cambodia[9] and no analyses of routine health system data to estimate the stillbirth rates. This means a gap in context-specific evidence to feed into policy prioritising interventions or resources towards further preventing stillbirths in the country.This study aimed to assess facility-level stillbirth rates over time and explore factors associated with stillbirths in the largest maternity referral hospital in Cambodia over a four-year period.

## Methods

### Study design

This is a retrospective cross-sectional study of singleton births at a national maternity care centre in Phnom Penh, Cambodia between January 2017 and December 2020 with a nested case-control component.

### Study setting

The study used data collected from the National Maternal and Child Health Centre (NMCHC) - a government tertiary level hospital under the Ministry of Health located in Phnom Penh, the capital of Cambodia. Phnom Penh has a population of around 2.3 million and constitutes 14% of the total population of Cambodia which in 2019 was estimated to be approximately 16 million people.[10] The NMCHC is one of the largest tertiary maternity referral hospitals in the country with around 7,000 births a year and is one of the main referral centres for high-risk cases. The NMCHC was built in 1997 and provides both clinical perinatal care and is a leading training institution in the country.[11] It has 134 in-patient beds, a neonatal care and an intensive care unit.[12]

### Study population

The study included all women who had a singleton birth at gestational age ≥22 weeks at the NMCHC between January 1, 2017, and December 31, 2020. For the cross-sectional analysis, we included all singleton births registered in the hospital database over the four-year period. For the case-control study, every singleton stillbirth over the four-year period was included and the consecutive singleton live birth after the stillbirth was selected as an unmatched control. Multiple births were excluded from the analysis.

### Data sources and data collection

Data on women and their baby were obtained directly from the NMCHC electronic hospital database of patients and additional data was extracted from individual medical records of women. For the case-control analysis, all stillbirths were identified from the hospital database and the consecutive singleton live birth was selected as an unmatched control. Individual medical files from each case and control were located and data extracted into a pre-structured form to cross-check information and provide additional data on variables which were not included in the hospital database.

Data on women’s obstetric history including parity, history of stillbirth or miscarriages/abortion, age, residence, gestational age, mode of birth, presentation of baby and birthweight of the baby were obtained from the electronic hospital database. Data on women’s past maternal medical conditions (hypertension/oedema, vaginal discharge or infections), history of caesarean birth, history of pre-term birth, and resuscitation of the index newborn were extracted from the individual women’s medical files.

### Study variables

The definition of stillbirth we applied is that outlined in ICD-11 and adopted by the WHO for early fetal deaths as a baby born with no signs of life from 22 weeks of pregnancy onwards.[4, 5] From the hospital database stillbirths were identified and confirmed based on the live status of baby indicated in the patient record sheet (from woman’s medical file) as either stillborn (*mort-né*) or alive (*vivant*), or if Apgar score was zero at 1, 5, and 10 minutes after birth in the medical file. Stillbirths were further categorised into intrapartum (fresh) or antepartum (macerated) using data from the electronic hospital database based on assessment the skin appearance recorded by the healthcare provider. In 2.1% of stillbirths, the timing was missing (20/938).

For the case-control study, the main outcome of interest was stillbirth (cases) and live births (controls). Independent variables included characteristics with a known relationship with the outcome of stillbirth according to published literature[7] and which were available in the hospital database and/or individual medical files. They included maternal demographic characteristics (maternal age and residence), obstetric history (parity, history of stillbirth/abortion/miscarriage, preterm birth or caesarean birth), fetal factors (gestational age, sex and birthweight, congenital malformation,) and obstetric factors (mode of birth, presentation of fetus, indication for caesarean section, resuscitation attempt) and history of maternal conditions (abnormal vaginal discharge or hypertension/oedema). Not all variables were included in the regression analysis; variables were selected after considering their relationship with the outcome and excluding those with a causal relationship or where timing of when the variable was measured or occurred after the outcome. The birthweight variable was re-categorised according to size for gestational age to create three categories – small for gestational age (SGA), normal for gestational age and large for gestational age. The details of how these were categorised are in the **Supplementary file (Table A1)**.

### Statistical analysis

Analyses were performed using STATA/SE v14 and SAS v9.4. For the cross-sectional analysis, we calculated the percentage of stillbirths among all births for each year and overall, and the stillbirth rates for each year as the number of stillbirths per 1000 total births. We also calculated these rates by timing of the stillbirth (intrapartum or antepartum) and the proportion of stillbirths by timing overall. Stillbirths with missing timing (20/938) were included in all calculations as a separate category. In the case-control study, we calculated descriptive statistics to summarise characteristics of women who had a stillbirth compared to those with a live birth. Chi-square tests was used to compare the percentages of cases and controls in terms of selected categorical variables. Bivariate analysis was performed using a binary logistic regression to examine the association between each independent variable and the outcome and to calculate unadjusted odds ratios with 95% confidence intervals. Independent variables which were statistically significant at p<0.25 in the bivariate analysis were retained and included in the multivariable logistic regression model. It was decided *a priori* to retain maternal age and baby’s sex in the models as it is established that either very young or older maternal age is an important risk factor for stillbirth and male babies are biologically at a slightly higher risk of stillbirth than females.[7]

Multivariable logistic regression was used to identify factors independently associated with the outcome. Two multivariable regression models were fitted – the first model included all cases and controls (n=1,876), while the second model included only women with at least one previous birth (n=1,046) so we could examine having had a previous pregnancy loss as a risk factor for stillbirth. We fit these models also on the full data set (with imputed values for observations missing gestational age and all variables relying on gestational age) and also on the data set with complete cases only to identify if the imputation affected or changed the overall results (see below) (**Supplementary file, Table A3**). In the multivariable models, independent variables that were not significant at 5% level were removed one at a time. To assess multi-collinearity, variance inflation factors were checked. Model fit was checked using the Hosmer-Lemeshow goodness of fit test. Due to the known relationship between birthweight and gestational age, we explored whether an interaction existed between these variables. Independent variables that were considered for the first multivariable logistic regression were maternal age, residence, gestational age, sex of the baby, hypertension/oedema, vaginal discharge, presentation of baby and mode of birth. The second multiple logistic regression added obstetric history factors such as history of stillbirth, abortion/miscarriages, and premature birth.

### Handling of missing values

Gestational age had the highest missingness with approximately 40% of observations among the controls (live births) and 48% of cases (stillbirths) missing gestational age in the case-control study dataset (n=1876). As we created a new variable of baby size at birth taking into account gestational age, this variable also had the same level of missingness. For the gestational age variable and size at birth, imputation was used with a total of 10 imputations to fill in missing values. These variables were first recoded into categories using available continuous values and this was followed by imputation of missing values as categorical variables. This approach was chosen over imputing gestational age as a continuous variable prior to categorising the variable, as it minimised variation in the estimates.

Missing data for other explanatory variables was <25% and these were almost all from observations from the control group (live births). Variables with missing data included hypertension/oedema (n=155/1876), history of vaginal discharge (n=110/1876), and history of premature birth (n=144/1876). As the number of observations with missing data in these variables was small, we recoded those observations with missing values to the category “No”.

## Results

### Levels and trends in stillbirth mortality at NMCHC

Of 29,742 singleton births in the 4-year period there were 938 stillbirths, giving an overall stillbirth rate of 32 per 1000 births or 3.2% of births (Table 1). Two-thirds (66%) of stillbirths were intrapartum while one-third occurred in the antepartum period. The stillbirth rate increased from 25 per 1000 births in 2017 to 38 per 1000 births in 2020 (Figure 1). When disaggregated by type of stillbirth, we found that the increase was due predominantly to an increase in intrapartum stillbirth rate in 2019 and particularly in 2020, whereas there was almost no change in the antepartum stillbirth rate.

**Table 1.**
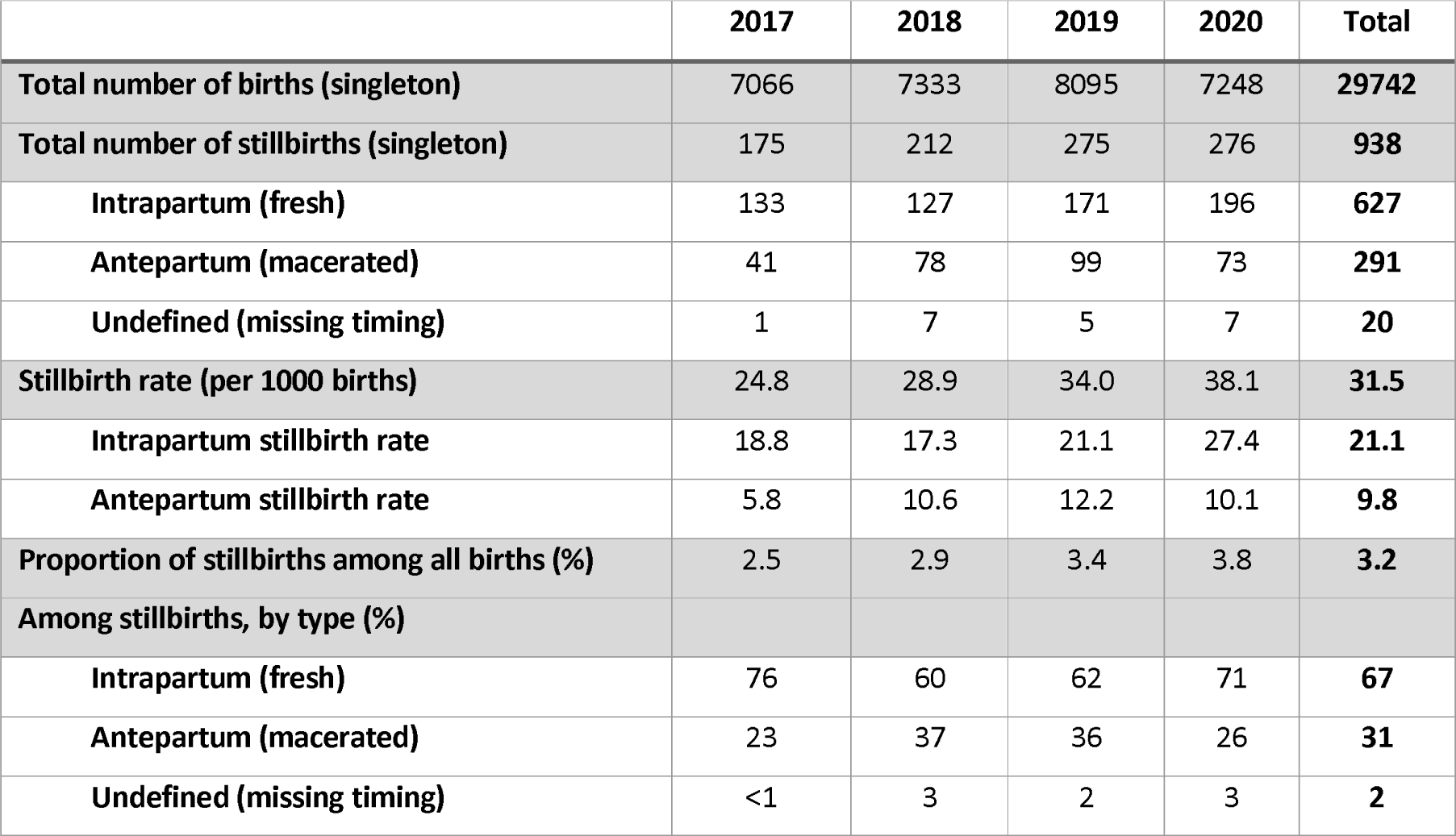
Proportion of stillbirths and stillbirth timing among singleton births, 2017-2020 at NMCHC, Cambodia.

|  | 2017 | 2018 | 2019 | 2020 | Total |
| --- | --- | --- | --- | --- | --- |
| **Total number of births (singleton)** | 7066 | 7333 | 8095 | 7248 | **29742** |
| **Total number of stillbirths (singleton)** | 175 | 212 | 275 | 276 | **938** |
| **Intrapartum (fresh)** | 133 | 127 | 171 | 196 | **627** |
| **Antepartum (macerated)** | 41 | 78 | 99 | 73 | **291** |
| **Undefined (missing timing)** | 1 | 7 | 5 | 7 | **20** |
| **Stillbirth rate (per 1000 births)** | 24.8 | 28.9 | 34.0 | 38.1 | **31.5** |
| **Intrapartum stillbirth rate** | 18.8 | 17.3 | 21.1 | 27.4 | **21.1** |
| **Antepartum stillbirth rate** | 5.8 | 10.6 | 12.2 | 10.1 | **9.8** |
| **Proportion of stillbirths among all births (%)** | 2.5 | 2.9 | 3.4 | 3.8 | **3.2** |
| **Among stillbirths, by type (%)** |  |  |  |  |  |
| **Intrapartum (fresh)** | 76 | 60 | 62 | 71 | **67** |
| **Antepartum (macerated)** | 23 | 37 | 36 | 26 | **31** |
| **Undefined (missing timing)** | <1 | 3 | 2 | 3 | **2** |

**Figure 1.**
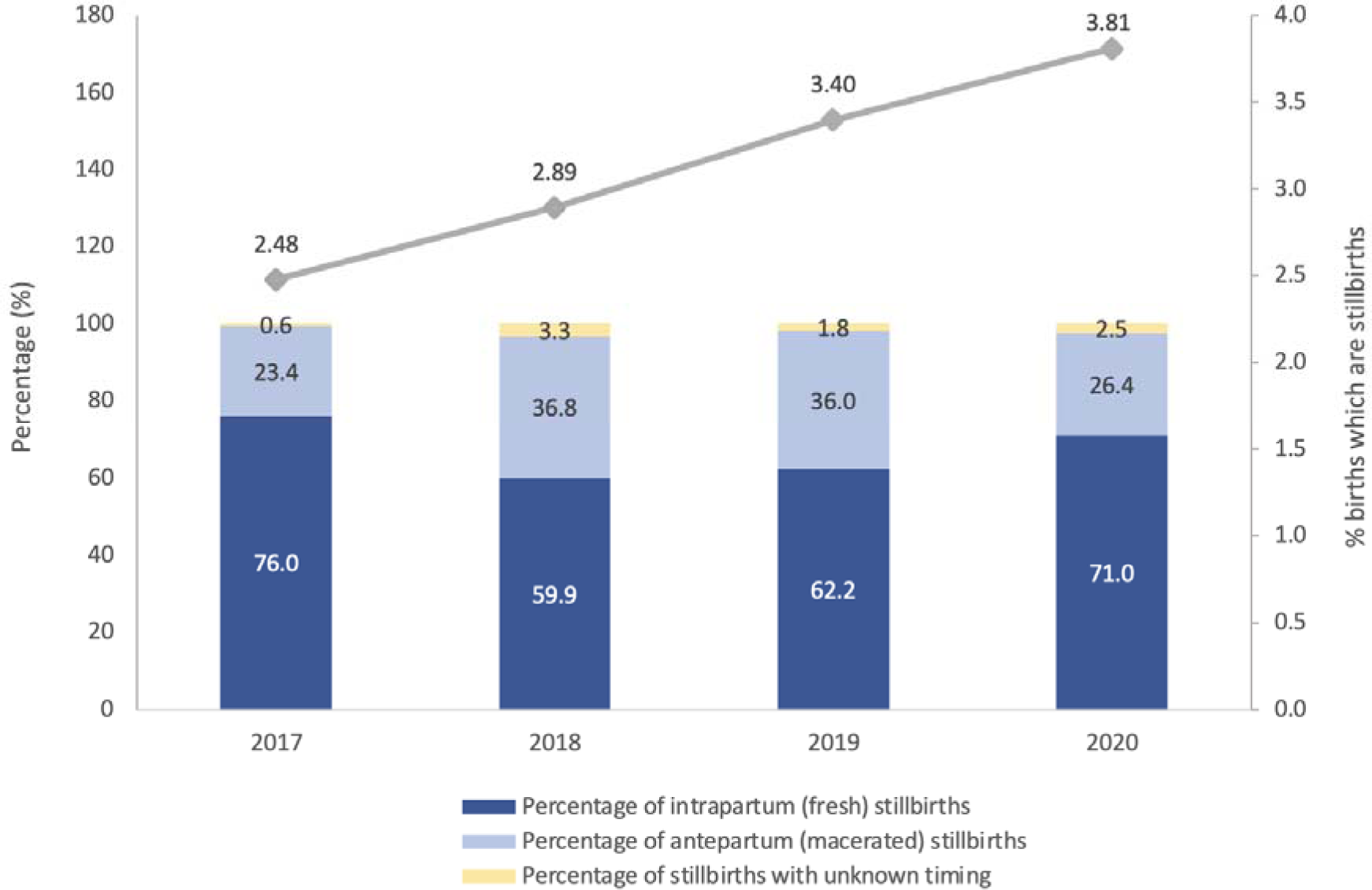
Proportion of stillbirths and timing of stillbirth between 2017-2020 among women with singleton births at NMCHC, Cambodia

### Case control study

#### Characteristics of cases and controls

There were 938 cases (stillbirths) and 938 controls (live births) included in the case-control analysis. Table 2 summarises the maternal, fetal and obstetric factors in the sample by outcome. Around 20% of women were over 35 years with only a small percentage <20 years. For just under half of the sample (44%) this was their first child, while about a quarter (25.3%) had two or more children. A history of at least one previous stillbirth was found in 2.5% of women; however, this was over double among women whose current birth ended in a stillborn (3.6%) compared to women who had a live birth (1.3%). A substantially higher percentage of live births were born at term or higher (37 or more weeks) while a higher percentage of stillborn babies were born extremely, very preterm, or moderate to late preterm. Overall, there were very few women giving birth after 41 weeks (5.6%). Among live births, over half were of normal weight for the gestational age but for stillborn babies almost 18% were small for gestational age. The rate of congenital malformations was more common among stillborn (6.0%) babies compared to liveborn (0.3%). Overall, 17% of babies were breech, transverse or face but the percentage was substantially higher among stillbirths (26%). The overall caesarean section (CS) rate was 27% but was lower among stillborn babies (21.4%) compared with live born (33.1%). Of note was that when we examined the indication for the CS in nearly 13% had fetal death given as the reason. Resuscitation was attempted on almost half of live born babies but only among 7% of stillborn babies. Among intrapartum stillbirths’ resuscitation was attempted on 9.1% (57/628) (results not shown). Around one-fifth of women had a history of abnormal vaginal discharge indicative of an infection, and this was slightly higher among women with a stillbirth (24.6%) compared to those whose baby was alive (19.4%). Only 2% of women had a recorded history of hypertension and this did not differ by the outcome.

**Table 2.** Characteristics of cases (stillbirths) and controls (live births), 2017-2020 NMCHC, Cambodia (n=1,876)

| Independent variable | Live birth |  | Stillbirth |  | Total |  |
| --- | --- | --- | --- | --- | --- | --- |
|  | n | % | n | % | n | % |
| *Maternal characteristics* |  |  |  |  |  |  |
| Maternal age (years) |  |  |  |  |  |  |
| <20 | 60 | 6.4 | 82 | 8.7 | 142 | 7.6 |
| 20-34 | 733 | 78.1 | 644 | 68.7 | 1,377 | 73.4 |
| 35+ | 145 | 15.5 | 212 | 22.6 | 357 | 19.0 |
| Residence |  |  |  |  |  |  |
| Urban | 690 | 73.6 | 637 | 67.9 | 1,327 | 70.7 |
| Semi-rural | 197 | 21.0 | 240 | 25.6 | 437 | 23.3 |
| Rural | 51 | 5.4 | 61 | 6.5 | 112 | 6.0 |
| Parity |  |  |  |  |  |  |
| Nulliparity | 421 | 44.9 | 409 | 43.6 | 830 | 44.2 |
| 1 | 304 | 32.4 | 268 | 28.6 | 572 | 30.5 |
| 2+ | 213 | 22.7 | 261 | 27.8 | 474 | 25.3 |
| History of stillbirth |  |  |  |  |  |  |
| No | 926 | 98.7 | 904 | 96.4 | 1,830 | 97.6 |
| Yes (1 or more) | 12 | 1.3 | 34 | 3.6 | 46 | 2.5 |
| History of miscarriage/abortion |  |  |  |  |  |  |
| No | 601 | 64.1 | 591 | 63.0 | 1,192 | 63.5 |
| Yes | 337 | 35.9 | 347 | 37.0 | 684 | 36.5 |
| History of premature birth |  |  |  |  |  |  |
| No | 930 | 99.2 | 927 | 98.8 | 1,857 | 99.0 |
| Yes | 8 | 0.9 | 11 | 1.2 | 19 | 1.0 |
| History of caesarean birth |  |  |  |  |  |  |
| No | 840 | 89.6 | 880 | 93.8 | 1,720 | 91.7 |
| Yes | 98 | 10.5 | 58 | 6.2 | 156 | 8.3 |
| *Fetal factors* |  |  |  |  |  |  |
| Gestational age |  |  |  |  |  |  |
| extremely preterm <28 weeks | 4 | 0.43 | 69 | 7.36 | 73 | 3.89 |
| very preterm 28-31 weeks | 22 | 2.35 | 134 | 14.29 | 156 | 8.32 |
| moderate to late preterm 32-36 weeks | 68 | 7.25 | 150 | 15.99 | 218 | 11.62 |
| term 37-40 weeks | 407 | 43.39 | 125 | 13.33 | 532 | 28.36 |
| 41+ weeks | 76 | 8.1 | 30 | 3.2 | 106 | 5.65 |
| missing | 361 | 38.49 | 430 | 45.84 | 791 | 42.16 |
| Gestational age |  |  |  |  |  |  |
| extremely or very preterm <32 weeks | 26 | 2.77 | 203 | 21.64 | 229 | 12.21 |
| moderate to late preterm 32-36 weeks | 68 | 7.25 | 150 | 15.99 | 218 | 11.62 |
| term >37 weeks | 483 | 51.49 | 155 | 16.52 | 638 | 34.01 |
| missing | 361 | 38.49 | 430 | 45.84 | 791 | 42.16 |
|  | n | % | n | % | n | % |
| Birthweight |  |  |  |  |  |  |
| <1500g | 27 | 2.88 | 334 | 35.81 | 361 | 19.24 |
| 1500-2499g | 136 | 14.5 | 323 | 34.43 | 459 | 24.5 |
| 2500-3999g | 748 | 79.7 | 251 | 26.76 | 999 | 53.3 |
| +4000g | 27 | 2.9 | 30 | 3.2 | 57 | 3.0 |
| Size at birth |  |  |  |  |  |  |
| Small for gestational age | 60 | 6.4 | 268 | 28.6 | 328 | 17.48 |
| Normal for gestational age | 487 | 51.92 | 203 | 21.6 | 690 | 36.78 |
| Large for gestational age | 30 | 3.2 | 37 | 3.9 | 67 | 3.57 |
| missing | 361 | 38.49 | 430 | 45.8 | 567 | 42.16 |
| Sex of baby |  |  |  |  |  |  |
| Female | 441 | 47.0 | 450 | 48.0 | 891 | 47.5 |
| Male | 497 | 53.0 | 488 | 52.0 | 985 | 52.5 |
| Congenital malformation |  |  |  |  |  |  |
| Yes | 3 | 0.3 | 110 | 11.7 | 113 | 6.0 |
| No | 935 | 99.7 | 828 | 88.3 | 1,763 | 94.0 |
| *Obstetric factors* |  |  |  |  |  |  |
| Presentation of baby |  |  |  |  |  |  |
| Vertex | 867 | 92.4 | 694 | 74.0 | 1,561 | 83.2 |
| Breech, transverse or face | 71 | 7.6 | 244 | 26.0 | 315 | 16.8 |
| Mode of birth |  |  |  |  |  |  |
| Vaginal | 608 | 64.8 | 718 | 76.6 | 1,326 | 70.7 |
| Caesarean | 310 | 33.1 | 201 | 21.4 | 511 | 27.2 |
| *Indication for CS was fetal death* | 0 | 0.0 | 26 | 12.9 | 26 | 5.1 |
| Vacuum assisted | 20 | 2.1 | 19 | 2.0 | 39 | 2.1 |
| Resuscitation attempted |  |  |  |  |  |  |
| Yes | 395 | 42.1 | 64 | 6.8 | 459 | 24.5 |
| No | 543 | 57.9 | 874 | 93.2 | 1,417 | 75.5 |
| *Maternal history of medical conditions* |  |  |  |  |  |  |
| Abnormal vaginal discharge |  |  |  |  |  |  |
| Yes | 182 | 19.4 | 231 | 24.6 | 413 | 22.0 |
| No | 756 | 80.6 | 707 | 75.4 | 1463 | 78.0 |
| Hypertension/oedema |  |  |  |  |  |  |
| Yes | 18 | 1.9 | 20 | 2.1 | 38 | 2.0 |
| No | 920 | 98.1 | 918 | 97.9 | 1,838 | 98.0 |

CS- caesarean section; GA- gestational age

The characteristics of cases and controls included in the sub-group analysis of women that had at least one previous birth is provided in the Supplementary material (Table A2).

### Bivariate and multivariable logistic regression analysis

Maternal age group, residence, parity, gestational age, birthweight, presentation, mode of birth, and vaginal discharge were significantly associated with stillbirth in bivariate analysis (**Table 3**, column 1). **Table 3** shows the first multivariable model fitted with gestational age, birthweight, and size at birth each separately (Model 1A-1C), a model with an interaction term between gestational age and birth size (Model 1D) and a model with gestational age and birth size included together as separate variables (Model 1E) The multivariable models fit on the complete cases only are presented in the Supplementary file (**Table A3**).

**Table 3.**
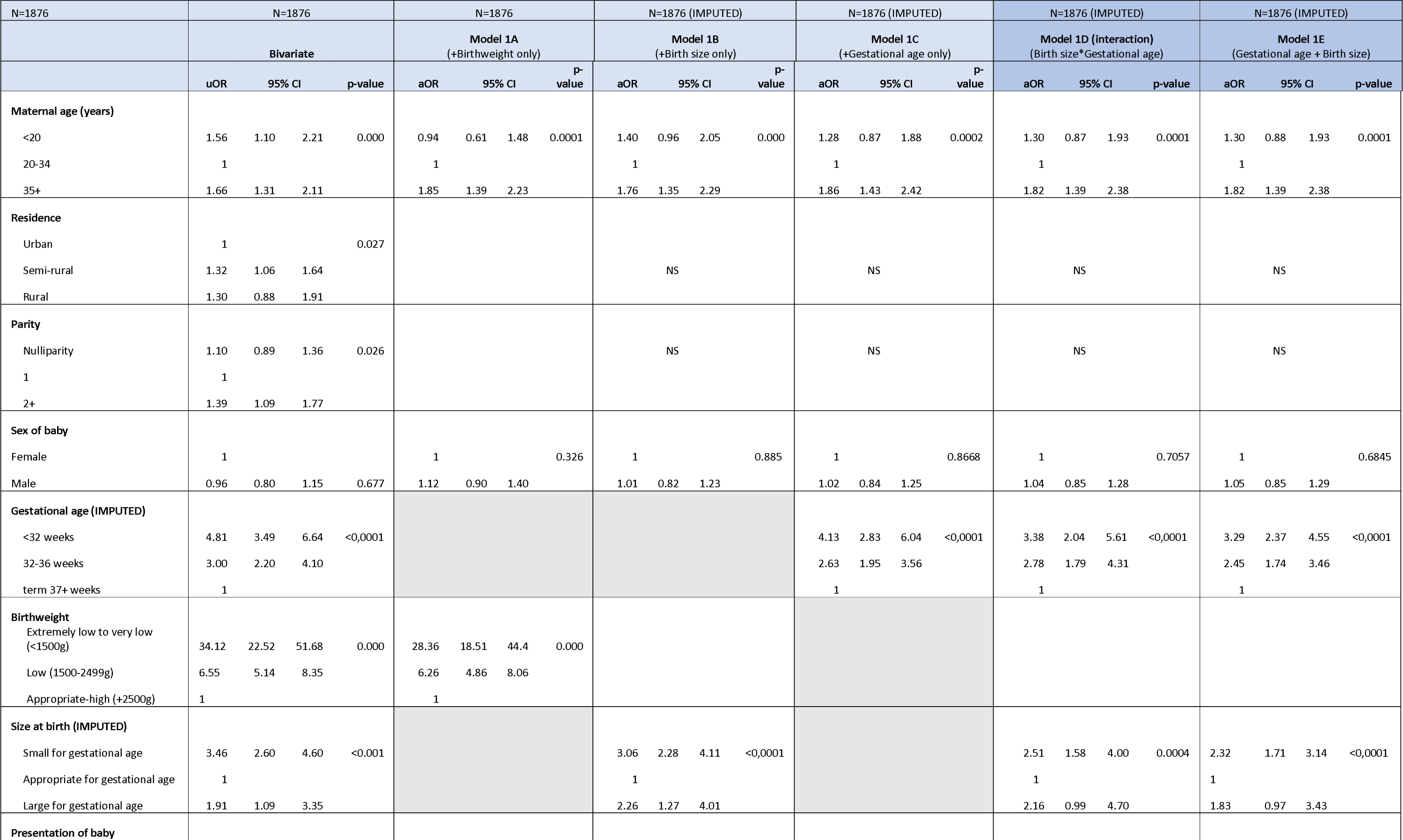
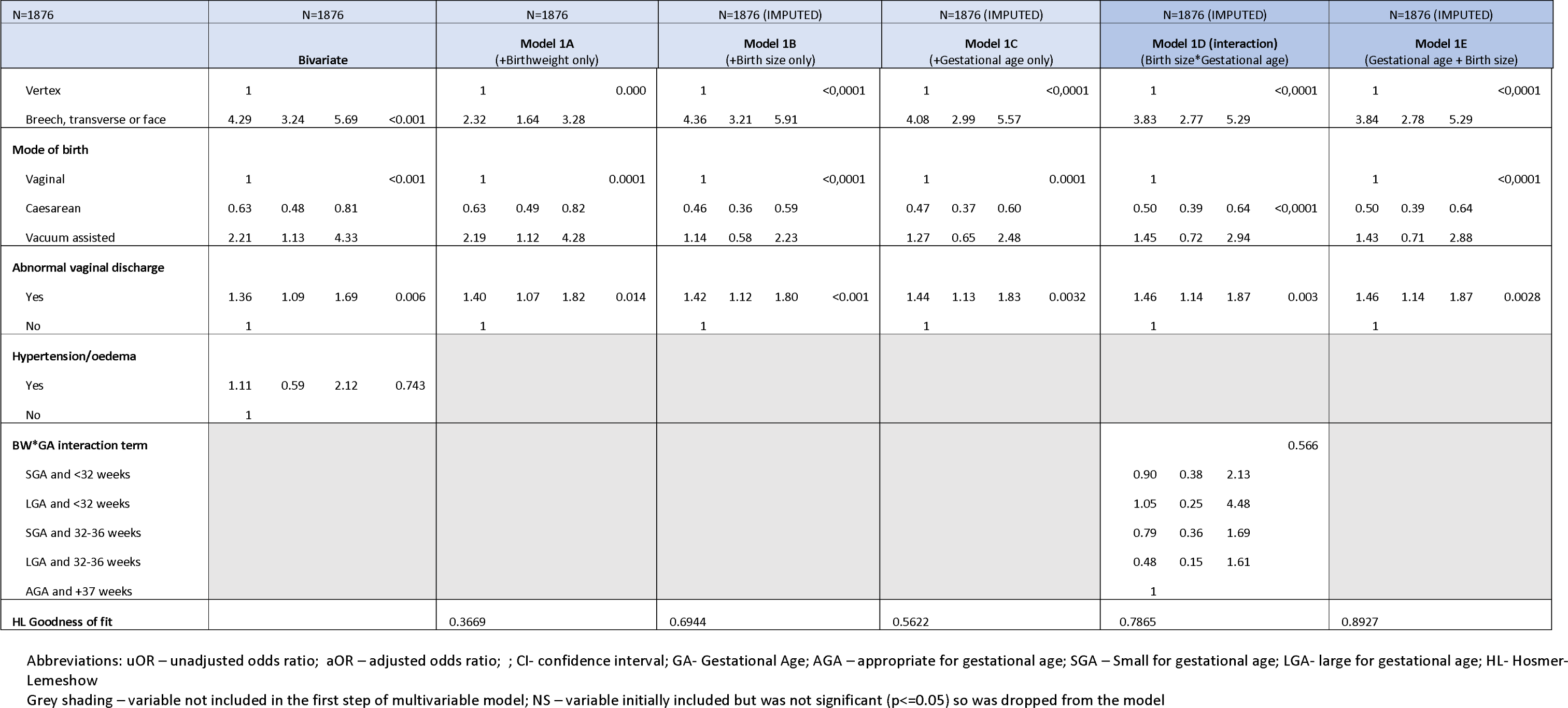
Bivariate and multivariable logistic regression of factors associated with stillbirth among all women (N=1876, Imputed values for women missing gestational age)

In the first multivariable model, maternal age was significantly associated with stillbirth; women aged >35 years had almost twice the odds of stillbirth (aOR 1.82, 95% CI 1.39-2.38) compared to women aged 20-34 years. Babies that were extremely premature (<32 weeks) had over three times increased odds of stillbirth and those moderately preterm had 2.5 times increase odds compared with babies born at term or higher. Babies that were small for gestational age or larger for gestational had 2.3- and 1.8-times greater odds of stillbirth, respectively. Babies that were born breech, transverse or face had almost 4 times increased odds of stillbirth compared to vertex presentation (aOR: 3.84, 95% CI: 2.78, 5.29). Babies born by caesarean section had half the odds of stillbirth compared to those born by vaginal birth (aOR 0.50, 95% CI: 0.39, 0.64). Baby’s sex and mother’s place of residence were not significantly associated with stillbirth after adjusting for other covariates (**Table 3**). There was no significant interaction between gestational age and birth size when the interaction was fit on the dataset with imputed values, however, in the complete case analysis the interaction was showing as significant (LR test p=0.03)(**Table A3**)

Model 2 (**Table 4**) shows the multivariable logistic regression results on the sub-group of women with at least one previous pregnancy. We found that a history of stillbirth increased the odds of stillbirth three-fold (aOR: 3.08, 95% CI: 1.48 6.43) compared to multipara without a history of stillbirth, after adjusting for other factors.

**Table 4.**
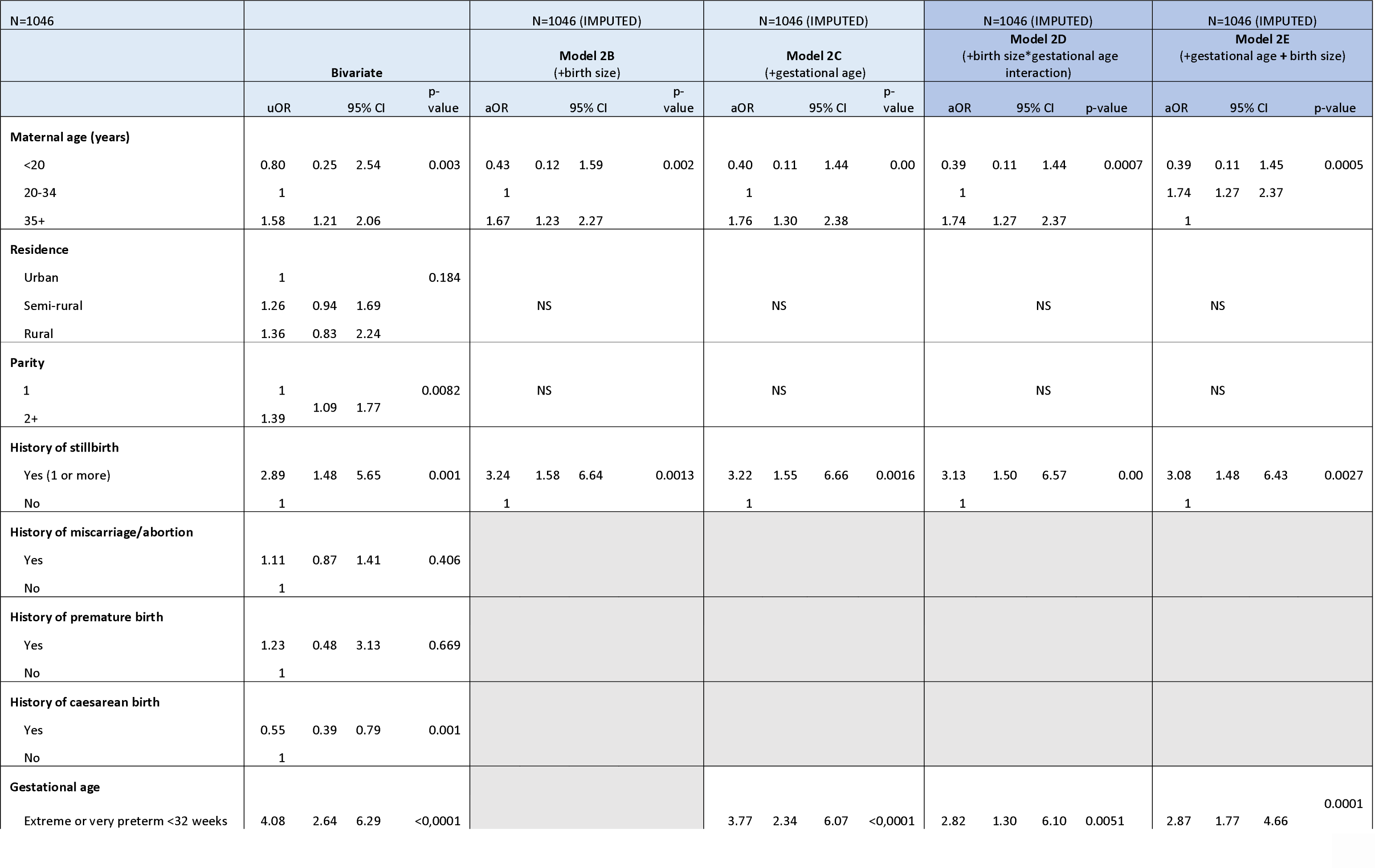
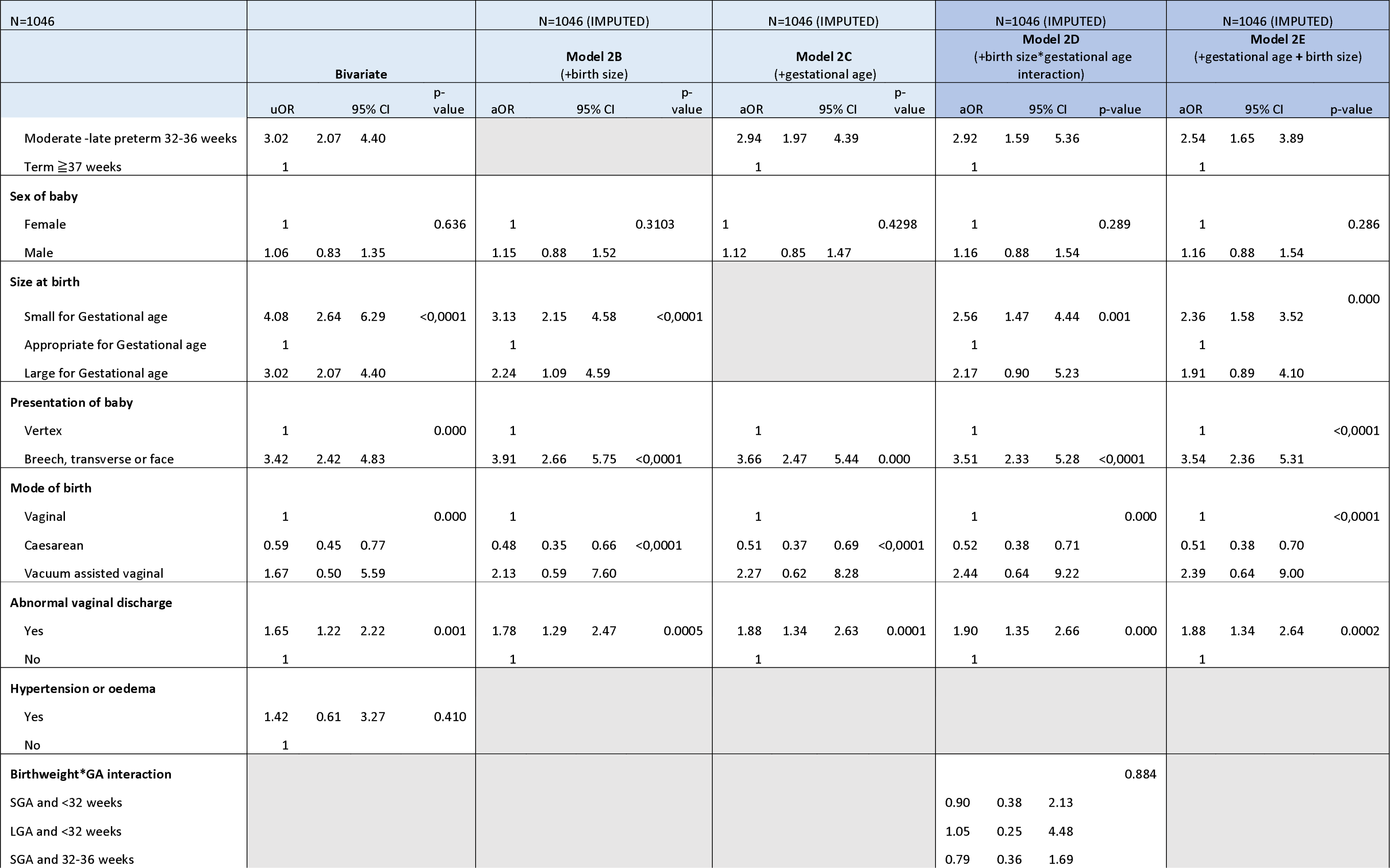
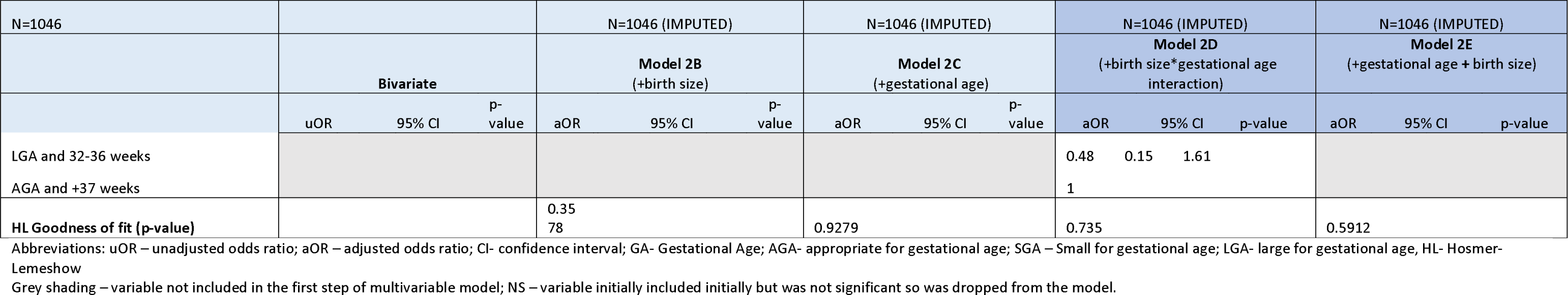
Multivariable logistic regression on sub-group of women with <u>at least one previous birth</u> (N=1046, Imputed values for women with missing gestational age)

## Discussion

This study is one of the first analyses using multi-year routine facility data on stillbirths in Cambodia from a high-volume public tertiary referral maternity hospital. We assessed the stillbirth burden and identified potential risk factors that can be targeted for prevention. Our study found an overall stillbirth rate at 32 per 1000 births among almost 30,000 births between 2017-2020. We found that the stillbirth rate in this hospital increased annually between 2017 and 2020 to a high of 38 per 1000 births in 2020 (60% higher than in 2017). Key factors that increased the odds of having a stillbirth included being aged over 35 years, having a baby that was born preterm or small for gestational age or with breech or transverse presentation, reporting abnormal vaginal discharge during pregnancy and a history of stillbirth. Giving birth by caesarean section was protective and reduced the odds of stillbirth.

The overall stillbirth rate at NMCHC was high compared to the national stillbirth (population) rates estimated for Cambodia which suggest a rate of 11 per 1000 births[13]. However, this is not unexpected given our analysis was from a tertiary referral maternity hospital and that this is not a population-based study representative of all births in Cambodia. A recent community-based study in Cambodia reported a stillbirth rate of 11 per 1000 births which aligns with levels in the national survey[9]. Facility-based stillbirth rates are commonly higher than population-based levels due to high risk or complicated births in (referral) maternities and case-mix. There are no other facility based studies within Cambodia to compare with, but in neighbouring Vietnam, facility-based stillbirth rates range from 25 per 1000 births in one tertiary facility in Ho Chi Minh city [14] to 9 per 1000 live births in Da Nang city where stillbirths from seven facilities were examined.[15].

The increase in the stillbirth rate over the four years could be the result of increased or better referral of high-risk cases as NMCHC is known to receive such cases from surrounding facilities and provinces. The peak in the intrapartum stillbirth in 2020 also coincides with the beginning of the COVID-19 pandemic. Although there were few COVID-19 cases in Cambodia until 2021, there were strict travel restrictions and school closures in place from early 2020[16], which could have led to delays for women to reach a health facility or less staff available, affecting both timely access to and ability to provide quality antenatal and emergency obstetric care. Of concern is that the increase in stillbirth rate over this four-year period was attributed to an increase in fresh or intrapartum stillbirth rate which indicates the baby is alive on arrival to the facility. Therefore, some of these deaths could potentially be saved if timely and quality obstetric care could be ensured. The share of intrapartum stillbirths in NMCHC accounted for 71% of all stillbirths in 2020, which is relatively high when the average estimated for low- and lower-middle-income countries is around 50% and for the East and South-East Asia region it is even lower at 30%. However, we acknowledge our measure of establishing the timing of stillbirth using skin appearance has its limitations.[17]

Our analysis identified several factors that were significantly associated with a higher odds of stillbirth in Cambodian mothers giving birth at NMCHC. The increased risk for women over 35 years and below 16 years of age is a known factor in the literature[7]. Our sample of women was too small to create an age group below 16 years, and we did not observe an increased risk of stillbirth in the youngest age group in our analysis (<20 years).

We found a significantly higher odds or stillbirth among babies who were extremely (<32 weeks) or moderately preterm (32-36 weeks), compared with term babies. There are limitations in interpreting this without additional knowledge on whether the baby was growing normally, as being born preterm could be the result of detection of FGR and therefore early induction or CS. When taking into account the baby’s birthweight for their gestational age, we found a significantly increased odds of stillbirth when the baby was either small for GA or large for GA. Several similar case-control studies that include a measure of GA also found high risk of stillbirth in preterm babies [18-20], and Recent modelling estimates suggest that 74% of stillbirths globally are among preterm babies, while a quarter of stillborn babies born at term were SGA[21]. Both preterm and SGA increase a babies’ vulnerability predisposing them to risk of both stillbirth and newborn death and longer-term consequences including stunting, disability, and non-communicable diseases.

Almost half of all babies in the case-control study were below 2500 grams – whether this is due to FGR or other factors is important to investigate for future stillbirth and neonatal mortality prevention in Cambodia, but also to improve longer term outcomes. Moreover, preterm, small for GA and stillbirth outcomes can also increase the risk of similar adverse outcomes in the next pregnancy so identifying and managing these risks will be critical to prevent future poor birth outcomes.[22]

Around a quarter of the stillborn babies in our study had a non-vertex presentation – the majority being breech. This led to almost four times greater risk of stillbirth compared with vertex presentation. Around two-thirds of breech babies were delivered vaginally in this facility, which warrants further examination on their management to understand why so many are leading to poor outcomes. Breech vaginal births can increase the risk of stillbirth and the current recommendation is to conduct CS for term breech presentation [23]. However, for pre-term breech births this remains contested, although CS appears to lower the risk of perinatal mortality in high income settings.[24] Nevertheless, consideration needs to be taken before recommending planned CS for breech presentations in low resource settings to avoid placing women or their baby at risk of additional complications due to the surgery and where there may be insufficient skills for CS.[25]

A history of stillbirth is a well-known risk factor for subsequent stillbirth which can increase the chance of stillbirth by four-fold or more[26]. It is therefore important to identify such women during their subsequent pregnancies to ensure closer monitoring and management to prevent recurrent stillbirths. Our study also showed an over three-fold increase in stillbirth among women who had a previous loss when we limited our analysis to women with a previous pregnancy. Many similar case-control analyses to ours do not exclude first time mothers from the sample used in the regression analysis which makes interpretation less reliable as these women have not had a previous pregnancy [20, 27, 28].

The overall CS rate in the case control sample was 27% and among stillbirths it was 21% and we found that having a CS was protective and reduced the chance of stillbirth by almost half. Among similar studies in the literature there is variation in terms of whether CS shows a protective effect[29] or increases stillbirth risk[18]. An increased risk could be attributable to a range of factors including delayed access to obstetric care or insufficient skills or resources to manage complications, as it is well established that CS reduces perinatal mortality when conducted appropriately in settings that are adequately resourced and skilled providers available[30].

One concerning finding regarding CS from our study was among the indications for CS for stillbirth we found that for almost 13% of stillbirths delivered by CS the indication documented was fetal death. Fetal death should not be a reason for CS unless there are other life-threatening complications to the mother. It is particularly the case if the stillbirth occurs in the antepartum period, as this places the woman at unnecessary risk of morbidity and further complications.[31] Additional inquiry into whether these CS were warranted may be required to ensure women are not placed at unnecessary risk.

We also found that among all stillbirths (intrapartum and antepartum) in this study, resuscitation attempts were very low compared with live births. Resuscitation has been shown to reduce the risk of both perinatal and neonatal mortality and current recommendations suggest that resuscitation should be attempted on every intrapartum stillborn baby.[4, 32] It is possible that resuscitation may have been attempted but was not recorded; however, this requires further exploration to ensure that no opportunity to save a baby is missed.

### Study strengths and limitations

Our study has several strengths – it presents one of the first published data on stillbirth in Cambodia, adding to the dearth of literature on this for the country. Moreover, we used routine data in a setting where over 90% of births occur in health facilities. We have a very large sample size over a four-year period from a large national maternity hospital which provides greater confidence in the findings and reducing the chance of errors. Our analysis also demonstrates the use of multiple imputation methods to address missing data on gestational age which can be applied to other similar routine data in LMIC. Missing data for gestational age is currently a major challenge in routine facility data in low resource settings[33] yet has important implications for stillbirth-for defining stillbirth and its use as a proxy for measuring FGR and preterm births.

Our study has several limitations in relation to the study design, variables available and the quality of routine data. This is single facility-based study and is therefore not representative at any geographic level. Being a referral facility, it is also likely to receive all high-risk cases which can bias the findings. We were unable to control for several important factors including socio-demographic determinants such as mother’s education and socio-economic status both of which are known risk factors for stillbirth. Our measure of mother’s residence as either urban or rural was also imprecise given the information available and be why we did not find any association as rural residence is commonly associated with increased stillbirth risk.

We had a large amount of missing data for some variables especially gestational age and although we addressed this using multiple imputation, this also has its limitations. Also because of this, we could not report on late gestation stillbirths from 28 weeks or more which is recommended for international comparisons.

Several other important factors were also not considered due to lack of or incomplete data or because the timing of the variable was unknown (e.g. for maternal conditions) including women’s antenatal care visits or content, referral status and use of obstetric interventions (e.g partograph) as well as clinical factors including maternal or fetal complications. We could not report directly on infections and used vaginal discharge as a proxy of possible infection although interpretation of this is limited and we have no knowledge on whether women received treatment for these or not. Syphilis and HIV testing are done routinely in Cambodia but there is no documentation of syphilis and HIV status at NMCHC [34] – these infections are important causes of stillbirth [7] and so need to be quantified and women treated as early as possible.

The distinction of stillbirth timing as fresh and macerated using assessment of skin condition can be inaccurate and so limits the interpretation of the proportions of antepartum and intrapartum stillbirths. Ideally, vital signs and fetal heart sounds on admission should used to differentiate between these[4], however we found that this data was of very low quality and inconsistent in routine records at NMCHC. There are various issues that can affect this data including available equipment to assess fetal heart sounds accurately and health provider skills in using these, as well as intentional misclassification to avoid blame.

## Conclusions and recommendations

Our study has quantified and identified factors associated with stillbirth in Cambodia using routine health facility data for the first time. Improved management of preterm births, and detection and management of SGA earlier in pregnancy as well as closer monitoring of women with history of stillbirth and aged over 35 years will be imperative for future stillbirth prevention in Cambodia. Further research is needed to understand the increase in stillbirths at this tertiary maternity hospital during 2017-20 and beyond and whether this is attributed to referrals of high-risk pregnancies, related to quality of care, or other reasons. A recent study in three provinces in Cambodia measuring knowledge of intrapartum care among skilled birth attendants found low levels of knowledge[35]. Review of the management of breech births in this facility is also needed. Future studies in Cambodia with a prospective design to capture all key factors and particularly infections such as HIV and syphilis and including a qualitative component can provide further data in the short-term. Strengthening the quality of facility data overall and gestational age should be emphasised to better inform future stillbirth prevention.

## Data Availability

The datasets generated and/or analysed during the current study are not publicly available as they are part of a hospital patient record database of the National Maternal and Child Health Centre in Cambodia. The authors do not have permission or approval to share this database beyond the investigators involved in the study.

## Declarations

### Ethics approval

The study has received ethical approval from the National Ethics Committee for Health Research in Cambodia (# 153/19; amendment # 294/20) and the Institutional Review Board at the Institute of Tropical Medicine, Belgium (#1317/19).

### Consent for publication

Not applicable

## Conflicts of interest

The authors declare they have no competing interests.

## Funding

This study received funding from the Belgian Directorate-General for Development Cooperation and Humanitarian aid (DGD). Aliki Christou is funded by the Research Foundation – Flanders (FWO) for a postdoctoral fellowship (number 1294322N). The funders were not involved in the study design; in the collection, analysis, interpretation of data; in the writing of the article; or in the decision to submit for publication.

## Author contributions

TD, MM, AI and KR conceptualised the study and secured funding. AC led the writing of the manuscript with critical comments provided by TD, LB, and MM. MM, TD, AN, SS, and NI oversaw data collection and data cleaning. AC and JM conducted the analysis with guidance from TD and LB. All authors have reviewed and approved the final version.

## List of Abbreviations

AGA: appropriate for gestational age
aOR: adjusted odds ratio
CI: confidence interval
CS: Caesarean section
FGR: fetal growth restriction
GA: Gestational age
HL: Hosmer Lemeshow
LGA: large for gestational age
LMIC: Low- and middle-income country
NMCHC: National Maternal and Child Health Centre
SGA: small for gestational age
uOR: unadjusted odds ratio
WHO: World Health Organisation

## Notes

### Competing Interest Statement

The authors have declared no competing interest.

### Author Declarations

The National Ethics Committee for Health Research in Cambodia (No. 153/19; amendment No.294/20) and the Institutional Review Board at the Institute of Tropical Medicine, Belgium (No.1317/19) gave ethical approval for this work.

## References

1. Hug L, You D, Blencowe H, Mishra A, Wang Z, Fix MJ, Wakefield J, Moran AC, Gaigbe-Togbe V, Suzuki E et al: Global, regional, and national estimates and trends in stillbirths from 2000 to 2019: a systematic assessment. The Lancet 2021, 398(10302):772–785.

2. UN IGME, UNICEF, WHO, World Bank, United Nations: Never Forgotten The situation of stillbirth around the globe. Report of the United Nations Interagency Group for Child Mortality Estimation, 2022. In. New York: UNICEF; 2023.

3. WHO: Every Newborn: an Action Plan to End Preventable Deaths. In. Geneva: WHO; 2014.

4. IGME, UNICEF, LSHTM: Stillbirth Definition and Data Quality Assessment for Health Management Information Systems (HMIS): A guideline. In. New York: UNICEF; 2022.

5. WHO: ICD-11 for Mortality and Morbidity Statistics. In.: WHO; 2023.

6. Pattinson R, Kerber K, Buchmann E, Friberg IK, Belizan M, Lansky S, Weissman E, Mathai M, Rudan I, Walker N et al: Stillbirths: how can health systems deliver for mothers and babies? Lancet 2011, 377(9777):1610-1623.

7. Lawn JE, Blencowe H, Waiswa P, Amouzou A, Mathers C, Hogan D, Flenady V, Froen JF, Qureshi ZU, Calderwood C et al: Stillbirths: rates, risk factors, and acceleration towards 2030. Lancet 2016, 387(10018):587–603.

8. National Institute of Statistics (NIS) Ministry of Health (MoH) Cambodia, ICF: Cambodia Demographic and Health Survey 2021–22 Key Indicators Report. In. Phnom Penh, Cambodia and Rockville, Maryland, USA: NIS, MoH, and ICF.; 2022.

9. Rambliere L, de Lauzanne A, Diouf J-B, Zo AZ, Landau M, Herindrainy P, Hivernaud D, Sarr FD, Sok T, Vray M: Stillbirths and neonatal mortality in LMICs: A community-based mother-infant cohort study. Journal of global health 2023, 13.

10. National Institute of Statistics MoP, Cambodia;: Report of Cambodia Socio-Economic Survey 2019/20. In. Phnom Penh: National Institute of Statistics, Ministry of Planning; 2020.

11. Koum K, Hy S, Tiv S, Sieng T, Obara H, Matsui M, Fujita N: Characteristics of antepartum and intrapartum eclampsia in the National Maternal and Child Health Center in Cambodia. Journal of Obstetrics and Gynaecology Research 2004, 30(2):74–79.

12. National Maternal and Child Health Center (NMCHC) Cambodia [https://nmchc.gov.kh/en/]

13. DHS program: Cambodia Demographic and Health Survey 2014. In.: National Insitute of Statisticas, Ministry of Planning and Ministry of Health; 2015.

14. Hirst JE, Arbuckle SM, Do TM, Ha LT, Jeffery HE: Epidemiology of stillbirth and strategies for its prevention in Vietnam. Int J Gynaecol Obstet 2010, 110(2):109–113.

15. Giang HTN, Bechtold-Dalla Pozza S, Tran HT, Ulrich S: Stillbirth and preterm birth and associated factors in one of the largest cities in central Vietnam. Acta paediatrica 2019, 108(4):630–636.

16. Chhim S, Ku G, Mao S, Put WVD, Damme WV, Ir P, Chhea C, Or V: Descriptive assessment of COVID-19 responses and lessons learnt in Cambodia, January 2020 to June 2022. BMJ Global Health 2023, 8(5):e011885.

17. Gold KJ, Abdul-Mumin AR, Boggs ME, Opare-Addo HS, Lieberman RW: Assessment of “fresh” versus “macerated” as accurate markers of time since intrauterine fetal demise in low-income countries. Int J Gynaecol Obstet 2014, 125(3):223–227.

18. Abebe H, Shitu S, Workye H, Mose A: Predictors of stillbirth among women who had given birth in Southern Ethiopia, 2020: A case-control study. PLOS ONE 2021, 16(5):e0249865.

19. Egbe TO, Ewane EN, Tendongfor N: Stillbirth rates and associated risk factors at the Buea and Limbe regional hospitals, Cameroon: a case-control study. BMC Pregnancy and Childbirth 2020, 20(1):75.

20. Okonofua FE, Ntoimo LFC, Ogu R, Galadanci H, Mohammed G, Adetoye D, Abe E, Okike O, Agholor K, Abdus-salam R et al: Prevalence and determinants of stillbirth in Nigerian referral hospitals: a multicentre study. BMC Pregnancy and Childbirth 2019, 19(1):533.

21. Lawn JE, Ohuma EO, Bradley E, Idueta LS, Hazel E, Okwaraji YB, Erchick DJ, Yargawa J, Katz J, Lee ACC et al: Small babies, big risks: global estimates of prevalence and mortality for vulnerable newborns to accelerate change and improve counting. The Lancet 2023, 401(10389):1707–1719.

22. Bane S, Simard JF, Wall-Wieler E, Butwick Alexander J, Carmichael SL: Subsequent risk of stillbirth, preterm birth, and small for gestational age: A cross-outcome analysis of adverse birth outcomes. Paediatr Perinat Epidemiol 2022, 36(6):815–823.

23. Hannah ME, Hannah WJ, Hewson SA, Hodnett ED, Saigal S, Willan AR: Planned caesarean section versus planned vaginal birth for breech presentation at term: a randomised multicentre trial. Term Breech Trial Collaborative Group. Lancet 2000, 356(9239):1375–1383.

24. Bergenhenegouwen L, Vlemmix F, Ensing S, Schaaf J, van der Post J, Abu-Hanna A, Ravelli ACJ, Mol BW, Kok M: Preterm Breech Presentation: A Comparison of Intended Vaginal and Intended Cesarean Delivery. Obstetrics and gynecology 2015, 126(6):1223–1230.

25. Sandall J, Tribe RM, Avery L, Mola G, Visser GHA, Homer CSE, Gibbons D, Kelly NM, Kennedy HP, Kidanto H et al: Short-term and long-term effects of caesarean section on the health of women and children. The Lancet 2018, 392(10155):1349–1357.

26. Lamont K, Scott NW, Jones GT, Bhattacharya S: Risk of recurrent stillbirth: systematic review and meta-analysis. Bmj 2015, 350:h3080.

27. Lema G, Mremi A, Amsi P, Pyuza JJ, Alloyce JP, McHome B, Mlay P: Placental pathology and maternal factors associated with stillbirth: An institutional based case-control study in Northern Tanzania. PLOS ONE 2021, 15(12):e0243455.

28. Tolefac PN, Tamambang RF, Yeika E, Mbwagbaw LT, Egbe TO: Ten years analysis of stillbirth in a tertiary hospital in sub-Sahara Africa: a case control study. BMC research notes 2017, 10:1–6.

29. Dagne HM, Melku AT, Abdi AA: Determinants of stillbirth among deliveries attended in Bale Zone Hospitals, Oromia Regional State, Southeast Ethiopia: a case–control study. International Journal of Women’s Health 2021, 13:51.

30. Darmstadt GL, Yakoob MY, Haws RA, Menezes EV, Soomro T, Bhutta ZA: Reducing stillbirths: interventions during labour. BMC Pregnancy Childbirth 2009, 9 Suppl 1(1):S6.

31. Zethof S, Christou A, Benova L, van Roosmalen J, van den Akker T: “Too much, too late”: data on stillbirths to improve interpretation of caesarean section rates. Bull World Health Organ 2022, 100(4):289–291.

32. Patel A, Khatib MN, Kurhe K, Bhargava S, Bang A: Impact of neonatal resuscitation trainings on neonatal and perinatal mortality: a systematic review and meta-analysis. BMJ Paediatr Open 2017, 1(1):e000183.

33. Miller L, Wanduru P, Santos N, Butrick E, Waiswa P, Otieno P, Walker D: Working with what you have: How the East Africa Preterm Birth Initiative used gestational age data from facility maternity registers. PLOS ONE 2020, 15(8):e0237656.

34. Delvaux T, Ouk V, Samreth S, Yos S, Tep R, Pall C, Keo V, Deng S, Khin Cho WH, Hul S et al: Challenges and outcomes of implementing a national syphilis follow-up system for the elimination of congenital syphilis in Cambodia: a mixed-methods study. BMJ Open 2023, 13(1):e063261.

35. Matsui M, Saito Y, Po R, Taing B, Nhek C, Tung R, Masaki Y, Iwamoto A: Knowledge on intrapartum care practices among skilled birth attendants in Cambodia—a cross-sectional study. Reproductive Health 2021, 18(1):115.

