## Supplementary file for "Stillbirth rates and their determinants in a national maternity hospital in Phnom Penh, Cambodia in 2017-2020: a cross-sectional assessment with a nested case-control study"

**Supplementary Appendix**

**Table A1.** Summary of variables considered among cases and controls (N= 1876)

| Variables | Definition in the primary data | Categorisation | Source | Number and % observations with missing or not recorded (N=1876) | Missing data handling | Included in Model 1 | Included in Model 2 (parity>0) |
| --- | --- | --- | --- | --- | --- | --- | --- |
| Maternal characteristics |  |  |  |  |  |  |  |
| Maternal age (years) | Maternal age at birth | <20 years  20-34 years  ≧35 years | Hospital database | 0 | n/a | Yes | Yes |
| Place of residence | Categorisation of province of residence according to local knowledge | Urban  Semi-rural  Rural | Hospital database | 0 | n/a | Yes | Yes |
| Obstetric history |  |  |  |  |  |  |  |
| Parity | Number of live births and stillbirths | 0  1  2+ | Hospital database | 0 | n/a | Yes | Yes |
| History of stillbirth | Any stillbirth previous to the index pregnancy | No  Yes (1 or more) | Hospital database | 170 (9.1%) | Recoded to ‘No’ | No | **Yes** |
| History of abortion or miscarriage | Any abortion or miscarriage previous to the index pregnancy | No  Yes (1 or more) | Hospital database + medical file | 110 (6.9%) | Recoded to ‘No’ | No | **Yes** |
| History of premature birth | Any history of previous premature birth | No  Yes (1 or more) | Medical file | 144 (7.7%) | Recoded to ‘No’ | No | **Yes** |
| History of caesarean birth | History of caesarean birth previous to the index pregnancy | No  Yes (1 or more) | Medical file | 227 (12.1%) | Recoded to ‘No’ | No | No (related to caesarean birth in mode of birth variable for index birth) |
| Maternal conditions |  |  |  |  |  |  |  |
| Hypertension or oedema | Hypertension or oedema | No  Yes | Medical file | 155 (8.4%) | Recoded to ‘No’ | Yes | Yes |
| Abnormal vaginal discharge | The woman had (abnormal) vaginal discharge | No  Yes | Medical file | 443 (23.6%) | Recoded to ‘No’ | Yes | Yes |
| Infections | Woman had any of the following: vaginal discharge or malaria or TB or STI or hepatitis or pyelonephritis.  (Excluded measles, rubella, varicella as>80% of women were positive) | No  Yes | Medical file | 110 (6.9%) | Recoded to ‘No’ | No (timing unknown) | No (timing unknown) |
| Fetal factocs |  |  |  |  |  |  |  |
| Gestational age | Gestational age in weeks | <32 weeks (Extremely or v preterm)  32-36 weeks (Moderate to late preterm)  ≥37 weeks (term or higher) | Hospital database | 791 (42.2%) | Imputed | Yes | Yes |
| Sex | Sex of the fetus | Female  Male | Hospital database | 0 | n/a | Yes | Yes |
| Birthweight | Birthweight of fetus in grams | <1500g (extreme or v low)  1500-2499g (low)  +2500 (normal-high) | Hospital database | 0 | n/a | No – used BW adjusted for GA | No – used BW adjusted for GA |
| Size at birth (according to gestational age) | SMALL for GA:  <32 weeks & <1500g  32-36weeks & <2000g  ≥37 weeks & <2500g  NORMAL for GA:  <32 weeks & 1500-1999g  32-36 weeks & 2000-2999g  ≥37weeks & 2500-3999g  LARGE for GA:  <32 weeks & ≥2000g  32-36 weeks & ≥3000g  ≥37 weeks & ≥4000g | Small for gestational age  Normal for gestational age  Large for gestational age | Hospital database | 791 (42.2%) | Imputed | Yes | Yes |
| Resuscitation attempted | If resuscitation was attempted on the baby | No  Yes | Medical files | 110 (19.7%) | Nothing | No | No |
| Malformation | Whether the baby had any congenital malformation | No  Yes | Hospital database | 0 | n/a | No- causal | No - causal |
| Obstetric factors |  |  |  |  |  |  |  |
| Mode of birth | Mode of birth | Normal vaginal  Assisted vaginal (vacuum)  Caesarean section | Hospital database | 0 | n/a | Yes | Yes |
| Presentation | Presentation of fetus during childbirth | No  Yes (Breech, transverse or face) | Hospital database | 0 | n/a | Yes | Yes |
| Indication for caesarean section | Includes placenta previa, eclampsia, fetal distress etc | Various | Hospital database | 0 | n/a | No -bias | No- bias |
| Childbirth complications | Level or extent of vaginal tearing or cervical rupture | Yes (Vaginal tearing level 1,2,3, cervical rupture)  No | Medical file | 1025 (54.6%) | Nothing | No -timing unknown, also many missing | No -timing unknown, also many missing |

GA – gestational age; n/a- not applicable, TB - tuberculosis, STI – sexually transmitted infection

**Table A2**. Characteristics of cases and controls among a sub-sample of women with a parity at least 1 (N=1046)

|  | **Live birth** | | **Stillbirth** | | **total** | |
| --- | --- | --- | --- | --- | --- | --- |
|  | n | % | n | % | n | % |
| *Maternal characteristics* |  |  |  |  |  |  |
| **Maternal age** |  |  |  |  |  |  |
| <20 | 7 | 1.4 | 5 | 1.0 | 12 | 1.2 |
| 20-34 | 379 | 73.3 | 339 | 64.1 | 718 | 68.6 |
| 35+ | 131 | 25.3 | 185 | 35.0 | 316 | 30.2 |
| **Residence** |  |  |  |  |  |  |
| Urban | 382 | 73.89 | 364 | 68.8 | 746 | 71.3 |
| Semi-rural | 105 | 20.31 | 126 | 23.8 | 231 | 22.1 |
| Rural | 30 | 5.8 | 39 | 7.4 | 69 | 6.6 |
| *Obstetric history* |  |  |  |  |  |  |
| **Parity** |  |  |  |  |  |  |
| Low parity(1-2) | 439 | 84.9 | 411 | 77.7 | 850 | 81.3 |
| High parity(3+) | 78 | 15.1 | 118 | 22.3 | 196 | 18.7 |
| **History of stillbirth** |  |  |  |  |  |  |
| No | 505 | 97.7 | 495 | 93.57 | 1,000 | 95.6 |
| Yes (1 or more) | 12 | 2.3 | 34 | 6.4 | 46 | 4.4 |
| **History of miscarriage/abortion** |  |  |  |  |  |  |
| No | 283 | 54.7 | 276 | 52.2 | 559 | 53.4 |
| Yes | 234 | 45.3 | 253 | 47.8 | 487 | 46.6 |
| **History of premature birth** |  |  |  |  |  |  |
| No | 509 | 98.5 | 519 | 98.1 | 1,028 | 98.3 |
| Yes (1 or more) | 8 | 1.6 | 10 | 1.9 | 18 | 1.7 |
| **History of caesarean birth** |  |  |  |  |  |  |
| No | 423 | 81.8 | 471 | 89.0 | 894 | 85.5 |
| Yes (one or more) | 94 | 18.2 | 58 | 11.0 | 152 | 14.5 |
| *Fetal factors* |  |  |  |  |  |  |
| **Gestational age** |  |  |  |  |  |  |
| extremely preterm <28 weeks | 3 | 0.58 | 34 | 6.43 | 37 | 3.54 |
| very preterm 28-31 weeks | 14 | 2.71 | 71 | 13.42 | 85 | 8.13 |
| Moderate-late preterm 32-36 weeks | 36 | 6.96 | 87 | 16.45 | 123 | 11.76 |
| term 37-40 weeks | 222 | 42.94 | 71 | 13.42 | 293 | 28.01 |
| 41+ weeks | 33 | 6.38 | 12 | 2.27 | 45 | 4.3 |
| missing | 209 | 40.43 | 254 | 48.02 | 463 | 44.26 |
| **Sex of baby** |  |  |  |  |  |  |
| Female | 247 | 47.8 | 245 | 46.3 | 492 | 47.0 |
| Male | 270 | 52.2 | 284 | 53.7 | 554 | 53.0 |
| **Birthweight** |  |  |  |  |  |  |
| <1500g | 16 | 3.1 | 185 | 35.0 | 210 | 19.2 |
| 1500-2499 | 64 | 12.4 | 174 | 32.9 | 238 | 22.8 |
| 2500g+ | 437 | 84.5 | 170 | 32.1 | 607 | 58.0 |
| **Size at birth** |  |  |  |  |  |  |
| Small for GA | 30 | 5.8 | 145 | 27.4 | 175 | 16.73 |
| Normal for GA | 258 | 49.9 | 107 | 20.2 | 365 | 34.89 |
| Large for GA | 20 | 3.87 | 23 | 4.4 | 43 | 4.11 |
| missing | 209 | 40.43 | 254 | 48.0 | 463 | 44.26 |
| **Congenital malformation** |  |  |  |  |  |  |
| No | 514 | 99.4 | 471 | 89.0 | 985 | 5.8 |
| Yes | 3 | 0.6 | 58 | 11.0 | 61 | 94.2 |
| *Obstetric factors* |  |  |  |  |  |  |
| **Presentation of baby** |  |  |  |  |  |  |
| Vertex | 466 | 90.1 | 385 | 72.8 | 851 | 81.4 |
| Breech, transverse or face | 51 | 9.9 | 144 | 27.2 | 195 | 18.6 |
| **Mode of birth** |  |  |  |  |  |  |
| Normal vaginal | 320 | 61.9 | 384 | 72.6 | 704 | 67.3 |
| Caesarean | 193 | 37.3 | 137 | 25.9 | 330 | 31.6 |
| *indication was fetal death* |  |  |  |  |  |  |
| Vacuum assisted | 4 | 0.8 | 8 | 1.5 | 12 | 1.2 |
| **Resuscitation attempted (AD)** |  |  |  |  |  |  |
| Yes | 221 | 42.8 | 35 | 6.6 | 256 | 24.5 |
| No | 296 | 57.3 | 494 | 93.4 | 790 | 75.5 |
| *Maternal history medical conditions* | |  |  |  |  |  |
| **Vaginal discharge (AD)** |  |  |  |  |  |  |
| No | 425 | 82.21 | 390 | 73.7 | 815 | 77.9 |
| Yes | 92 | 17.8 | 139 | 26.28 | 231 | 22.1 |
| **Hypertension/oedema (AD)** |  |  |  |  |  |  |
| Yes | 9 | 1.74 | 15 | 2.8 | 24 | 2.3 |
| No | 508 | 98.3 | 514 | 97.2 | 1,022 | 97.7 |

**Table A3.** Bivariate and multivariable logistic regression of factors associated with stillbirth among complete cases only (N=1085, women without values for GA were excluded)

|  |  | | | | N=1876 (COMPLETE CASES) | | | | N=1085 (COMPLETE CASES) | | | | N=1085 (COMPLETE CASES) | | | | N=1085 (COMPLETE CASES) | | | | | | N=1085 (COMPLETE CASES) | | | |
| --- | --- | --- | --- | --- | --- | --- | --- | --- | --- | --- | --- | --- | --- | --- | --- | --- | --- | --- | --- | --- | --- | --- | --- | --- | --- | --- |
|  | **Bivariate** | | | | **Model 1A** (+ birthweight) | | | | **Model 1B**  (+birth size) | | | | **Model 1C** (+gestational age) | | | | **Model 1D** (birth size*GA interaction) | | | | | | **Model 1E** (Gestational age + birth size) | | | |
|  | **uOR** | **95% CI** | | **p-value** | **aOR** | **95% CI** | | **p-value** | **aOR** | **95% CI** | | **p-value** | **aOR** | **95% CI** | | **p-value** | **aOR** | | **95% CI** | | **p-value** | | **aOR** | **95% CI** | | **p-value** |
| **Maternal age (years)** |  |  |  |  |  |  |  |  |  |  |  |  |  |  |  |  |  | |  |  |  | |  |  |  |  |
| <20 | 1.56 | 1.10 | 2.21 | <0.001 | 0.94 | 0.61 | 1.44 | 0.0001 | 1.43 | 0.79 | 2.59 | 0.0001 | 1.01 | 0.54 | 1.87 | <0.001 | 0.99 | | 0.52 | 1.88 | 0.000 | | 1.06 | 0.56 | 1.99 | <0.0001 |
| 20-34 | 1 |  |  |  | 1 |  |  |  | 1 |  |  |  |  |  |  |  | 1 | |  |  |  | | 1 |  |  |  |
| 35+ | 1.66 | 1.31 | 2.11 |  | 1.85 | 1.39 | 2.47 |  | 2.30 | 1.56 | 3.39 |  | 2.49 | 1.67 | 3.72 |  | 2.62 | | 1.71 | 3.99 |  | | 2.53 | 1.67 | 3.83 |  |
| **Residence** |  |  |  |  |  |  |  |  |  |  |  |  |  |  |  |  |  | |  |  |  | |  |  |  |  |
| Urban | 1 |  |  | 0.027 |  |  |  |  |  |  |  |  |  |  |  |  |  | |  |  |  | |  |  |  |  |
| Semi-rural | 1.32 | 1.06 | 1.64 |  |  |  | NS |  |  | NS |  |  |  | NS |  |  |  | | NS |  |  | |  | NS |  |  |
| Rural | 1.30 | 0.88 | 1.91 |  |  |  |  |  |  |  |  |  |  |  |  |  |  | |  |  |  | |  |  |  |  |
| **Parity** |  |  |  |  |  |  |  |  |  |  |  |  |  |  |  |  |  | |  |  |  | |  |  |  |  |
| Nulliparity | 1.10 | 0.89 | 1.36 | 0.026 |  |  | NS |  |  | NS |  |  |  | NS |  |  |  | | NS |  |  | |  | NS |  |  |
| 1 | 1 |  |  |  |  |  |  |  |  |  |  |  |  |  |  |  |  | |  |  |  | |  |  |  |  |
| 2+ | 1.39 | 1.09 | 1.77 |  |  |  |  |  |  |  |  |  |  |  |  |  |  | |  |  |  | |  |  |  |  |
| **Gestational age** |  |  |  |  |  |  |  |  |  |  |  |  |  |  |  |  |  | |  |  |  | |  |  |  |  |
| <32 weeks | 24.33 | 15.57 | 38.03 | <0.001 |  |  |  |  |  |  |  |  | 21.39 | 13.36 | 34.24 | <0.001 | 25.81 | | 10.77 | 61.86 | <0.0001 | | 11.23 | 6.78 | 18.58 | <0.001 |
| 32-36 weeks | 6.87 | 4.90 | 9.65 |  |  |  |  |  |  |  |  |  | 6.97 | 4.86 | 10.02 |  | 6.29 | | 3.91 | 10.12 |  | | 4.97 | 3.39 | 7.27 |  |
| term >=37 weeks | 1 |  |  |  |  |  |  |  |  |  |  |  | 1 |  |  |  | 1 | |  |  |  | | 1 |  |  |  |
| **Sex of baby** |  |  |  |  |  |  |  |  |  |  |  |  |  |  |  |  |  | |  |  |  | |  |  |  |  |
| Female | 1 |  |  | 0.677 | 1 |  |  | 0.326 | 1 |  |  | 0.496 | 1 |  |  | 0.278 | 1 | |  |  | 0.318 | | 1 |  |  | 0.218 |
| Male | 0.96 | 0.80 | 1.15 |  | 1.12 | 0.90 | 1.40 |  | 1.10 | 0.83 | 1.47 |  | 1.18 | 0.87 | 1.60 |  | 1.17 | | 0.86 | 1.61 |  | | 1.22 | 0.89 | 1.66 |  |
| **Birthweight** |  |  |  |  |  |  |  |  |  |  |  |  |  |  |  |  |  | |  |  |  | |  |  |  |  |
| Extremely low to very low (<1500g) |  |  |  |  | 28.36 | 18.51 | 43.44 | <0.001 |  |  |  |  |  |  |  |  |  | |  |  |  | |  |  |  |  |
| Low (1500-2499g) |  |  |  |  | 6.26 | 4.86 | 8.06 |  |  |  |  |  |  |  |  |  |  | |  |  |  | |  |  |  |  |
| Appropriate to high (+2500g) |  |  |  |  | 1 |  |  |  |  |  |  |  |  |  |  |  |  | |  |  |  | |  |  |  |  |
| **Size at birth (adjusted for GA)** |  |  |  |  |  |  |  |  |  |  |  |  |  |  |  |  |  | |  |  |  | |  |  |  |  |
| Small for GA | 10.63 | 8.42 | 13.42 | <0.001 |  |  |  |  | 9.26 | 6.57 | 13.05 | 0.000 |  |  |  |  | 6.52 | | 3.74 | 11.37 | 0.000 | | 4.12 | 2.80 | 6.06 | 0.000 |
| Appropriatel for GA | 1 |  |  |  |  |  |  |  | 1 |  |  |  |  |  |  |  | 1 | |  |  |  | | 1 |  |  |  |
| Large for GA | 3.47 | 2.24 | 5.38 |  |  |  |  |  | 4.18 | 2.42 | 7.20 |  |  |  |  |  | 3.59 | | 1.57 | 8.20 |  | | 2.18 | 1.17 | 4.06 |  |
| **Presentation of baby** |  |  |  |  |  |  |  |  |  |  |  |  |  |  |  |  |  | |  |  |  | |  |  |  |  |
| Vertex | 1 |  |  |  | 1 |  |  | <0.001 | 1 |  |  | 0.000 | 1. |  |  | 0.000 | 1 | |  |  | 0.000 | | 1 |  |  | 0.000 |
| Breech, transverse or face | 4.29 | 3.24 | 5.69 | <0.001 | 2.32 | 1.64 | 3.28 |  | 4.50 | 2.88 | 7.03 |  | 3.36 | 2.09 | 5.40 |  | 2.70 | | 1.64 | 4.45 |  | | 2.77 | 1.69 | 4.52 |  |
| **Mode of birth** |  |  |  |  |  |  |  |  |  |  |  |  |  |  |  |  |  | |  |  |  | |  |  |  |  |
| Vaginal | 1 |  |  | <0.001 | 1 |  |  | 0.0001 | 1 |  |  | 0.0000 | 1 |  |  | 0.0000 | 1 | |  |  | 0.0000 | | 1 |  |  | 0.00 |
| Caesarean | 0.55 | 0.45 | 0.68 |  | 0.63 | 0.49 | 0.81 |  | 0.36 | 0.26 | 0.52 |  | 0.46 | 0.32 | 0.66 |  | 0.45 | | 0.31 | 0.66 |  | | 0.47 | 0.32 | 0.68 |  |
| Vacuum assisted vaginal | 0.80 | 0.43 | 1.52 |  | 2.19 | 1.12 | 4.28 |  | 2.10 | 0.92 | 4.80 |  | 2.90 | 1.26 | 6.66 |  | 3.90 | | 1.66 | 9.16 |  | | 3.57 | 1.53 | 8.30 |  |
| **Abnormal vaginal discharge** |  |  |  |  |  |  |  |  |  |  |  |  |  |  |  |  |  | |  |  |  | |  |  |  |  |
| Yes | 1.36 | 1.09 | 1.69 | 0.006 | 1.40 | 1.07 | 1.82 | 0.014 | 1.55 | 1.10 | 2.19 | 0.013 | 1.77 | 1.24 | 2.55 | 0.002 | 1.72 | | 1.18 | 2.52 | 0.005 | | 1.73 | 1.19 | 2.51 | 0.004 |
| No | 1 |  |  |  | 1 |  |  |  | 1 |  |  |  | 1 |  |  |  | 1 | |  |  |  | | 1 |  |  |  |
| **Hypertension/oedema** |  |  |  |  |  |  |  |  |  |  |  |  |  |  |  |  |  | |  |  |  | |  |  |  |  |
| Yes | 1.11 | 0.59 | 2.12 | 0.743 |  |  |  |  |  |  |  |  |  |  |  |  |  | |  |  |  | |  |  |  |  |
| No | 1 |  |  |  |  |  |  |  |  |  |  |  |  |  |  |  |  | |  |  |  | |  |  |  |  |
| **Birth size*GA (Interaction)** |  |  |  |  |  |  |  |  |  |  |  |  |  |  |  |  |  | | | | | |  |  |  |  |
| SGA <32 weeks |  |  |  |  |  |  |  |  |  |  |  |  |  |  |  |  | 0.20 | 0.06 | | 0.63 | | 0.03 |  |  |  |  |
| LGA and <32 weeks |  |  |  |  |  |  |  |  |  |  |  |  |  |  |  |  | 0.54 | 0.22 | | 1.30 | |  |  |  |  |  |
| SGA and 32-36 weeks |  |  |  |  |  |  |  |  |  |  |  |  |  |  |  |  | 0.31 | 0.06 | | 1.71 | |  |  |  |  |  |
| LGA and 32-36 weeks |  |  |  |  |  |  |  |  |  |  |  |  |  |  |  |  | 0.28 | 0.07 | | 1.08 | |  |  |  |  |  |
| AGA and +37 weeks |  |  |  |  |  |  |  |  |  |  |  |  |  |  |  |  | 1 |  | |  | |  |  |  |  |  |
| **HL Goodness of fit** |  |  |  |  |  |  |  |  | 0.4051 |  |  |  | 0.8874 |  |  |  | 0.8088 | |  |  |  | | 0.057 |  |  |  |
| **LR test** |  |  |  |  |  |  |  |  |  |  |  |  |  |  |  |  | **LR test p=0.0261** | | | | | | | |  |  |

Grey shading – variable not included in the first step of multivariable model; NS – variable initially included initially but was not significant so was dropped from the model

aOR – adjusted odds ratio, uOR – unadjusted odds ratio, CI- confidence interval, AGA – Appropriate for Gestational Age, SGA – Small for gestational age, LGA- large for gestational age, LR – Likelihoood Ratio Test
